# Beyond Exploratory Data: Using Cluster Detection Tests to Pinpoint Malaria Hotspots in Nigeria

**DOI:** 10.1101/2024.06.04.24308429

**Authors:** S. O. Oyamakin, Owolabi Mayowa

## Abstract

Malaria is not communicable, but it can deteriorate and even kill a person within a day if treatment is not taken. Plasmodium falciparum is the malaria parasite that is most prevalent and deadly in Africa. This study attempts to identify malaria hotspots and potentially high-risk areas in Oyo state, Nigeria. Data on malaria cases from 2013 to 2016 were provided by the Department of Research, Planning, and Statistics (DPRS) of the Oyo State Ministry of Health. Spatial autocorrelation testing, spatiotemporal analysis, and descriptive statistics were performed using SatScan V9.11 and ArcGIS V10.8. The four-year malaria incidence study was conducted in Oyo West LGA and is geographically focused. Four clusters were found using spatial analysis, with the majority of the clusters located in the Oyo South region. Spatial-temporal research revealed three noteworthy groups. The cluster that happened between January 2013 and December 2014 is located in the Oyo South and Oyo Central region. High-high malaria clusters were identified by spatial autocorrelation analysis, particularly in Oyo Central; the yearly clustered pattern of malaria incidence was not coincidental. In order to better achieve effective disease surveillance, this study suggests employing more cluster detection tests (or CDTs) rather than just exploratory data analysis prior to the implementation of health treatments and regulations.

## Introduction

The parasites that cause malaria include Plasmodium falciparum and Plasmodium vivax, which bite humans and create a potentially fatal sickness. The deadliest malaria parasite and most common in Africa is Plasmodium falciparum, which poses the most threat. Outside of sub-Saharan Africa, Plasmodium vivax is the predominant malaria parasite. Within 24 hours, P. falciparum malaria can progress to severe illness and death if left untreated. The early symptoms of malaria, which include fever, headache, and chills, usually appear 10 to 15 days after the infective mosquito bite and can be mild or hard to diagnose. Malaria posed a threat to about half of the world's population in 2020. Infants, children under five, pregnant women, people living with HIV/AIDS, and people with low immunity who are traveling to areas where malaria transmission is intense, such as migrant workers, mobile populations, and travelers, are among the population groups that have a significantly higher risk of contracting malaria and developing severe disease (World Malaria report, 2021).

**Figure 1:**
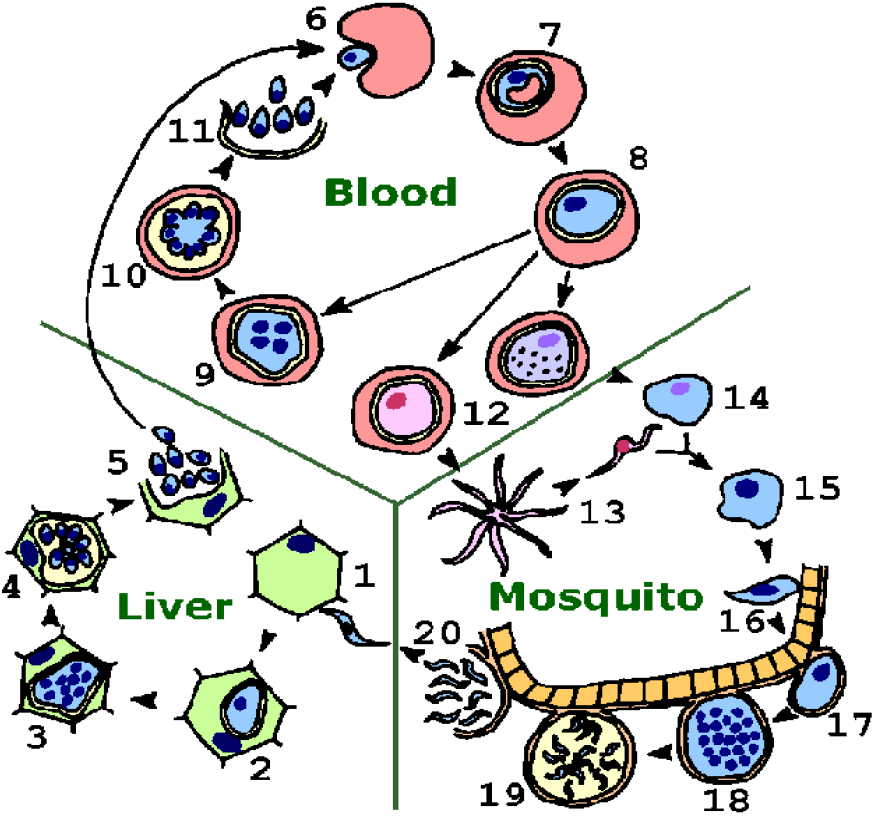
Transmission cycle of Malaria. Note. The image in figure 1 was sourced from Tulane University website. https://www2.tulane.edu/~wiser/protozoology/notes/images/mal_lc.gif. The image in figure 2 was sourced from Krekora et al. (2017); the course of pregnancy and delivery in a patient with malaria. Ginekologia Polska. 88. 574-575. 10.5603/GP.a2017.0103.

**Figure 2:**
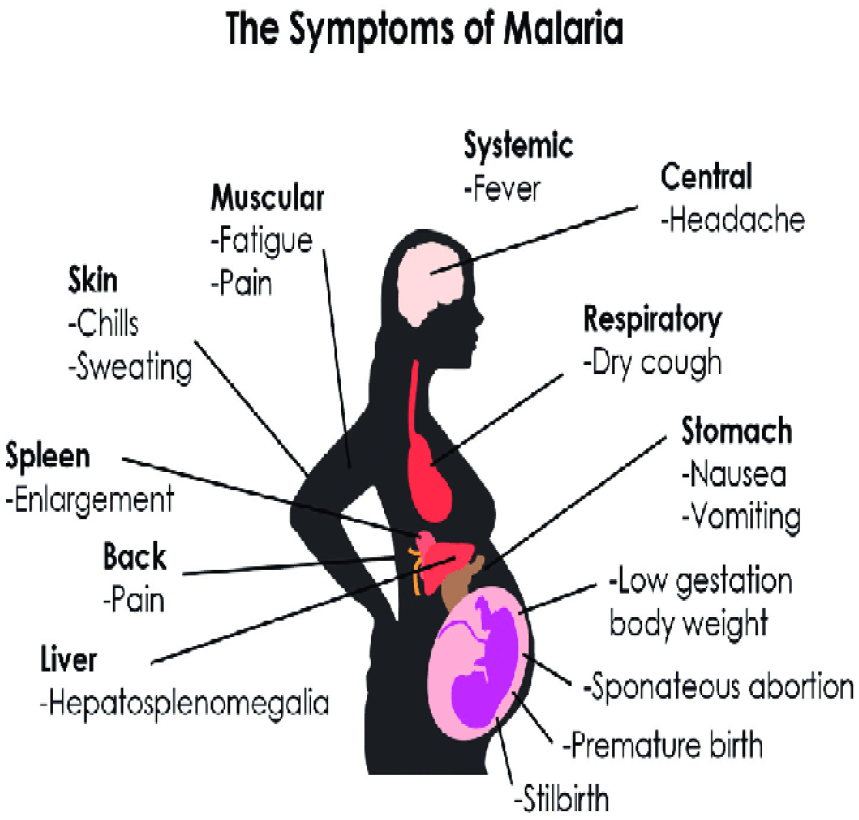
Signs and Symptoms of Malaria

Oyo State, popularly referred to as the “Pace Setter,” is the fifth most populous state in Nigeria and one of the 36 states that comprise the Federal Republic of Nigeria. With a total area of roughly 28,454 square kilometers, it is the fourteenth largest state in Nigeria in terms of size. Oyo State shares borders with Ogun State to the south, Kwara State to the north, Ogun State and the Republic of Benin to the west, and Osun State to the east. Oyo State has an equatorial climate with two distinct seasons: the dry season, which runs from November to March, and the wet season, which begins in April and finishes in October. There is also a fair amount of humidity in the state. The Oyos, Ogbomoshos, Oke-Oguns, Ibadans, and Ibarapas comprise the majority of its local population. They are all members of the Yoruba family and are indigenous to the region in southern Africa. Nigeria's Oyo State is home to 29 Local Council Development Areas and 33 Local Governments. The state's climate is ideal for the growth of crops such cashews, cocoa trees, palm trees, plantains, yam, cassava, rice, and plantains (oyostate.gov.ng).

**Figure 3:**
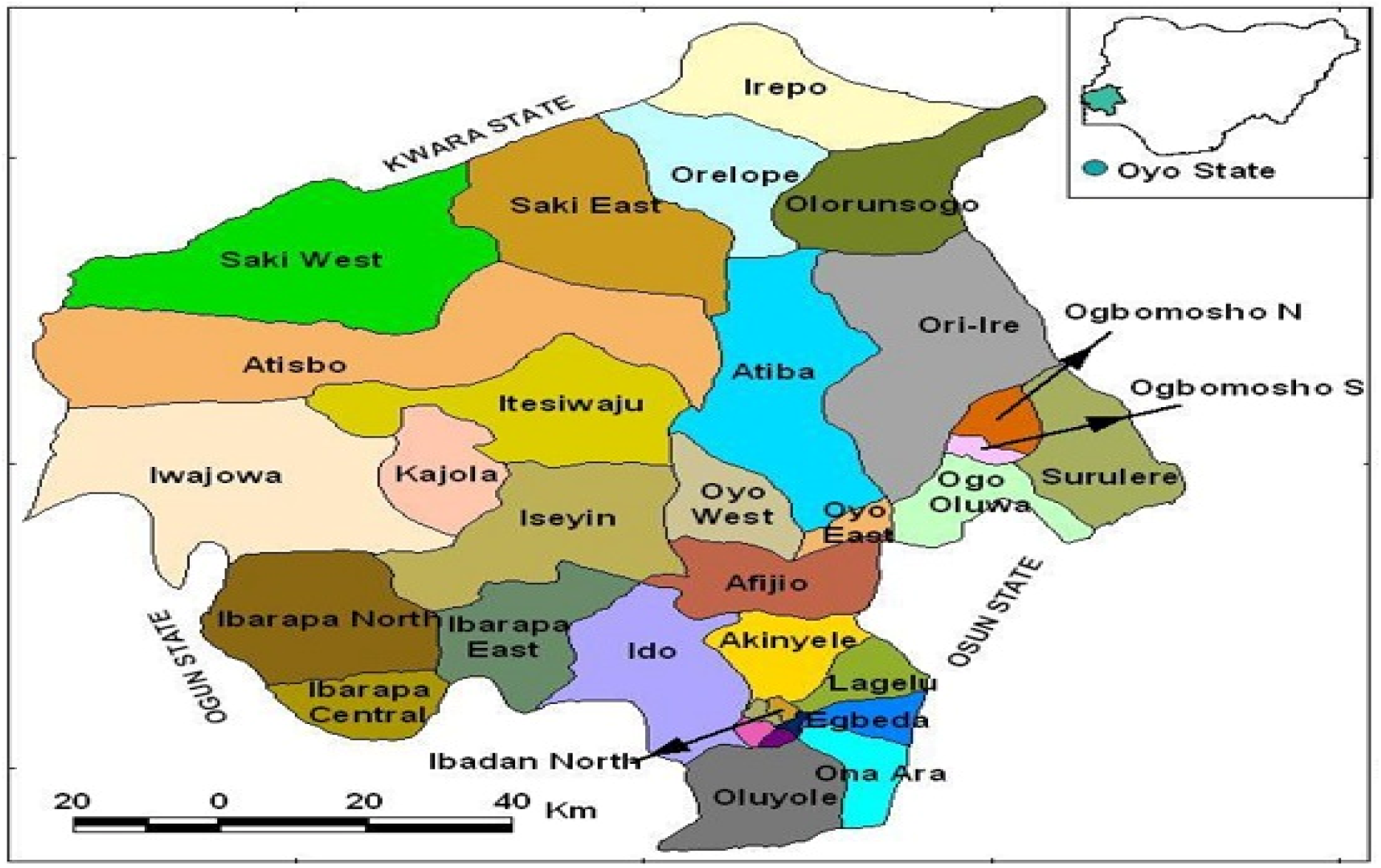
Geographical map of Oyo state. *Note*. A geographical map showing the thirty-three local governments in Oyo state, Nigeria.

Estimates indicate that 76% of Nigerians reside in regions where malaria transmission is high. Nigeria had the greatest percentage of malaria-related deaths (24%), accounting for 27% of all cases reported worldwide. Four African countries—Nigeria (31.9%), the Democratic Republic of the Congo (13.2%), Tanzania (4.1%), and Mozambique (3.8%)—accounted for more than half of all malaria-related deaths globally, according to the World Malaria Report 2020. The U.S. Malaria Initiative for States (PMI-S) was originally adopted by eight states in 2021: Akwa Ibom, Benue, Cross River, Ebonyi, Nasarawa, Oyo, Plateau, and Zamfara. The goal of its implementation was to reduce the mortality rates of mothers and children under five, as well as to improve treatment efficacy and accessibility to malaria services at all levels (USAID, 2021). Oyo State's membership in the previously mentioned project indicates that the state still has a high fatality rate from malaria. As a result, Nigeria needs more illness surveillance in order to provide evidence-based treatments. This comprehensive data would facilitate the distribution of health resources based on risk regions and enhance the availability of malaria treatment and prevention. The global community has paid close attention to the confluence of health crises, including the Novel Coronavirus Disease 2019 (COVID 19), cancer, tuberculosis, HIV/AIDS, and malaria, among others. Numerous tests have been developed to ascertain whether clustering is real and meaningful for such purposes.

Several statistical techniques have been used in disease surveillance to identify spatial clusters. Among these techniques, a subclass known as the generic test searches for clusters without speculating on their potential locations. But whether the statistical significance data for every cluster is accessible will depend on the method employed (Takahashi and Shimadzu, 2021). Developed by Moran (1950), Whitemore et al. (1987), Oden (1995), Tango (1995), Rogerson (1999), and Bonetti and Pagano (2005), global clustering tests (GCTs) are methods that do not ascertain the statistical significance of clusters. On the other hand, some techniques—such as those published by Turnbull et al. (1990), Besag and Newell (1991), Kulldorf and Nagarwalla (1995), Kulldorf (1997), and Tango (2000)—are referred to as cluster detection tests (or CDTs). Thus, there's a need to go over some of the study's supporting literature. Dead bird clusters were evaluated by Farzad et al. (2003) as a potential early warning system for West Nile Virus activity. The prospective detection of infectious disease outbreaks on dead birds reported from New York City in 2000 was accomplished using the spatial scan statistic. Its efficacy in offering a preemptive alert regarding West Nile Virus activity in 2001 was also examined. The study suggests that the development of an early warning system for West Nile virus (WNV) outbreaks could provide the basis for more rapid suppression of adult and larval mosquito populations, as well as more focused public awareness campaigns and monitoring activities. Furthermore, a prospective geographic cluster analysis of bird death reports may alert authorities to increased viral activity in birds and mosquitoes early on, allowing them to better allocate their limited laboratory and mosquito-collection resources and prevent human illness caused by the virus. They believed that additional infectious disease surveillance systems, such as those for bioterrorism, could benefit from the adaptation of the scan statistic.

Before and after adjusting for risk factors, Klassen et al. (2005) looked at the geographic clustering of prostate cancer grade and stage at diagnosis. The study combined projected block group-level disease trends from multilevel models with a spatial scan statistic technique to examine regional variance in prostate cancer grade and stage at diagnosis. According to the study's findings, Maryland prostate cancer cases between 1992 and 1997 fell into four statistically significant clusters with high and low rates of each outcome (later stage at diagnosis and higher histologic grade of tumor). This was accurate even prior to the modifications. After accounting for specific case factors like age, race, and year of diagnosis, the clustering patterns of both outcomes shifted. Before and after adjusting for risk factors, Klassen et al. (2005) looked at the geographic clustering of prostate cancer grade and stage at diagnosis. The study combined projected block group-level disease trends from multilevel models with a spatial scan statistic technique to examine regional variance in prostate cancer grade and stage at diagnosis. According to the study's findings, Maryland prostate cancer cases between 1992 and 1997 fell into four statistically significant clusters with high and low rates of each outcome (later stage at diagnosis and higher histologic grade of tumor). This was accurate even prior to the modifications. After accounting for specific case factors like age, race, and year of diagnosis, the clustering patterns of both outcomes shifted.

Fukuda et al. (2005) examined socioeconomic differences among geographic disease clusters using colon, lung, and breast cancer as case studies from Japan. In order to find spatial clusters of the three common types of cancer (breast, lung, and colon), as well as to identify dimensional socioeconomic factors and minimize the number of variables, the study used the spatial scan statistic and Principal Component Analysis (PCA), which was modified for seven socio-economic indicators. Using Japanese municipal data (N = 3360) from 1993 to 1998, the spatial scan statistic was used to identify spatial clusters of high mortality rates for female breast cancer and male colon and lung cancer. In addition, four sociological clusters (clutter, suburban, rural-poor, and urban-rich) were identified within municipalities based on population density and socioeconomic factors.

## METHODOLOGY

Because it works with geographic data, the study of spatial statistics is also known as spatial analysis. Spatial statistics is used in many different fields, and the methods employed depend on the particular research questions being addressed. Examples of these include managing the massive amounts of data generated by GPS (global positioning system) and satellite remote sensing, predicting soil quality in agriculture, and investigating unusually high frequencies of disease events. Spatial data is special because geographic position provides a key that is shared, either exactly or roughly, across data sets from different origins. There are three main types of spatial data: area, lines, and points. Points are features that have a specific location and no further reach in any direction. Cities, industrial zones, and towns are a few instances of point data. Line features are sequences of discrete x, y coordinate pairs with discrete beginning and ending points. Features include things like lines, rivers, and road networks. Shapes known as polygons are created by enclosing an area with a network of connecting lines. Polygons are defined by their area and perimeter. Administrative borders, land use, soil maps, etc. are a few instances of polygon characteristics (Prachi Sahoo, 2012).

## DESCRIPTIVE SPATIAL STATISTICS

### 1 Mean Center

The mean is a critical measure of central tendency for a set of data. When the concept of central tendency is applied to locational point data in two dimensions (X and Y coordinates), the mean center, sometimes referred to as the average location, can be found. It can be found by separately averaging the X and Y

coordinates, as follows:

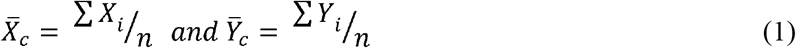

Where;
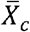 = mean center of X

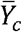 = mean center of Y

*X*_*i*_= X coordinate of point i

*y*_*i*_= Y coordinate of point i

n = number of points in the distribution

In many geographical applications, it is appropriate to assign different weights to the points in a spatial distribution. The weights are similar to the frequencies in the analysis of grouped data (weighted mean, etc.).

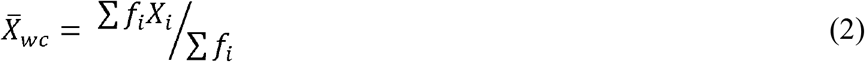

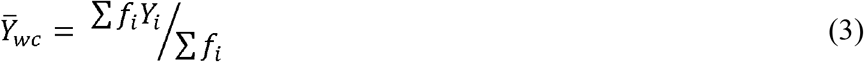

Where; 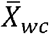 = weighted mean center of X, 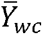 = weighted mean center of X, and *f*_*i*_= frequency (weight) of point i

## 2 Euclidean Median

The Euclidean median is an additional useful metric for determining the center. It is more practical to determine the center that minimizes the total unsquared distances rather than the total squared distances. The location that minimizes the total of the Euclidean distances from every other point in a geographical distribution to that central location is called the Euclidean median ( *X*_*e*_, *Y*_*e*_), or the median center. In this position, the sum is mathematically reduced:also known as the median center, is the place that minimizes the sum of Euclidean distances from all other points in a spatial distribution to that central location. This position reduces the sum mathematically:

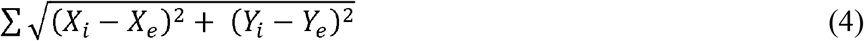

The simple (unweighted) Euclidean median is logically expanded to become a weighted Euclidean median. Population, sales volume, or any other feature suited to the spatial problem may be represented by the weights or frequencies. The weighted Euclidean median's (*X*_*we*_,*y*_*we*_) coordinates will minimize the expression.

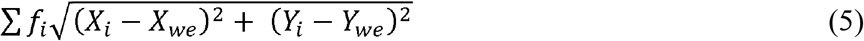

## 3 Standard Distance

A point pattern's absolute dispersion is measured using the standard distance (*S*_*D*_). The standard distance statistic includes the straight-line or Euclidean distance of each point from the mean center after the geographic coordinates of the mean center have been established. Following are some estimates for standard distance:

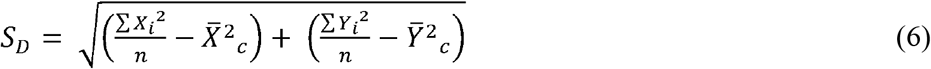

Weighted standard distance (*S*_*WD*_) is appropriate for those geographic applications requiring a weighted mean center.

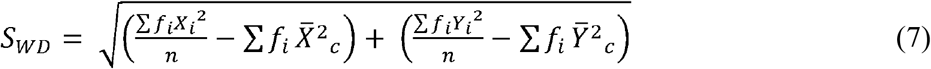

## 4 Spatio-Temporal Analysis - Spatial Scan Statistics

The spatial scan statistic, which is a generalization of the cluster evaluation permutation technique (CEPP) and is a spatial variant of the scan statistic with a variable window size, was proposed by Kulldorff and Nagarwalla (1995) and Kulldorff (1997). Each centroid of a region is given a circular window Z by the spatial scan statistic. The radius of the circle changes from zero to a predetermined upper limit for each of those centroids. The entire region is included in the window if it contains the centroid of an area. In total, a very large number of different but overlapping circular windows are created, each with a different location and size, and each being a potential cluster. Let *Z*_*ik*_,*K = 1*,*…, K*_*i*_, denote the window composed by the (*k −* 1) nearest neighbors to region *i*. Then, all the windows to be scanned by the spatial scan statistic are included in the set

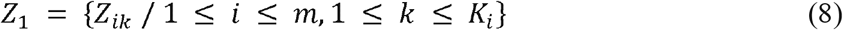

Under the alternative hypothesis, there is an elevated risk within some window **Z** as compared to outside:

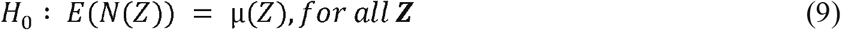

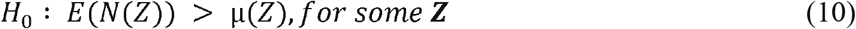

N() and μ() stand for the random number of cases and the expected number of cases, respectively, within the given window. The likelihood to observe the observed number of cases inside and outside of each window can be calculated separately. The most likely cluster (MLC) is identified by the window Z^*^ that maximizes the likelihood ratio. With the help of SaTScan Software, Kulldorff's spatial scan statistic has been used in a wide range of epidemiological investigations as well as disease surveillance for the detection of disease clusters (Kulldorff et al., 2007).

Spatial scan statistics can be applied to both aggregated data and data with specific point locations. The latter will be considered in the proposed study. There are m cells in a research area, C sickness cases overall, and N people in total. The null hypothesis states that we assume each cell's population size-dependent expected values are Poisson distributed and that there are no clusters in the map. We define L(Z) as the likelihood under the null hypothesis that there is no cluster in the zone and L_0 as the probability under the alternative hypothesis that there is a cluster in zone Z, per Duczmal and colleagues (2005).

If μ(Z) is the expected number of cases inside the zone Z under the null hypothesis, N is the number of observed cases during the entire study period; and μ(T) the total number of expected cases in the study area across all time periods (Desjardins et al. 2020). A likelihood ratio test is then used to identify clusters using;

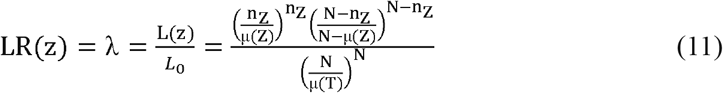

Where;
**λ** = Test statistic

**L(Z)** = Likelihood under the alternative hypothesis

*L*_0_ = Likelihood under the null hypothesis

**n**_**z**_ = The observed number of malaria cases inside the zone Z

**μ(Z)** = The expected number of malaria cases inside the zone Z

**μ(T) =** The total number of expected cases of malaria in the zone Z across all time periods

**N** = The number of observed malaria cases during the entire study period

Once the value of the test statistic has been calculated, it is easy to do the inference. If a zone exhibits a likelihood ratio greater than 1 i.e. 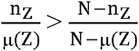, the risk is elevated. The zone that makes up the most likely cluster is identified by this likelihood ratio test, which maximizes over all of the zones (Kulldorf 1997). The likelihood ratio test, whose P-value is produced by using Monte Carlo simulations, is the foundation for the test of the importance of the detected clusters (Dwass 1957). We might wish to take a closer look at secondary clusters with high likelihood values in addition to the most likely cluster. These are defined as clusters that do not overlap with one that is more likely. Reporting these clusters along with the most likely one is frequently of interest. After accounting for demographic factors like age or race, we might also check to see whether any geographic clusters remain. This could point us in the direction of additional spatially associated risk variables that are currently unknown (Kulldorf, 1997).

## 5 Spatial Autocorrelation

The results of spatial autocorrelation show whether or not the observed value of a variable at one place is independent of its values at other locations. Correlation statistics were created to show interactions between variables, as opposed to autocorrelation statistics, which are intended to show correlations within variables (Artur, 2007). The link between variables across spatial dimensions is shown by spatial autocorrelation. The spatial statistical methods of global spatial autocorrelation and local spatial autocorrelation are used to characterize the relationship between the research regions and quantify the degree of aggregation or dispersion. The Moran's I index is a method for measuring spatial autocorrelation. While the global Moran's I index is used to evaluate the overall spatial autocorrelation and spatial distribution of the study areas, the local Moran's I index may also be used to reflect the local spatial autocorrelation and the particular clustering areas (Tille et al., 2018).

Moran's I index values vary from -1 to + 1. A positive autocorrelation is denoted by an I > 0, and the distribution of cases is aggregated spatially. A negative autocorrelation is shown by an I < 0, and the more examples are dispersed, the closer the value is to − 1. According to Mao et al. (2019), an I = 0 denotes a random distribution of the cases in space.

### Classical Measure of Spatial Autocorrelation

- Moran's I
- Geary's C

### 1 Global Moran I

**Moran (1950)** proposed a measure to estimate the global spatial autocorrelation (β);

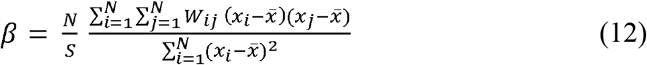

Where *X*_*i*_ is the observed value at location i, N is the number of locations and, 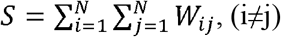, (i≠j). The weighting function *W*_*ij*_ is used to assign weights to every pair of locations in the study area, *W*_*ij*_ = 1, if I and j are neighbours, otherwise *W*_*ij*_ = 0.

### 2 Local Morans I

Local morans I (LISA, Anselin, 1995) is used to explore events that are clustered across geographic units. Local morans I is given below, where 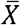 and *S*^2^ is the mean value and variance respectively.

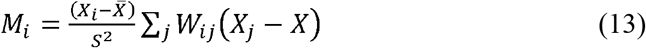

### 3 Geary's C

Moran's I provides a more general indication, but the Geary coefficient is more sensitive to changes in a small neighborhood. Geary's C does not offer the same inference since it highlights the values that differ between observational pairs rather than their covariation.

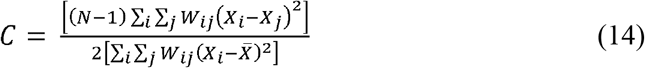

## DISCUSSION OF RESULTS

This section attempts to investigate the malaria prevalence in the 33 local governments of Oyo State by discovering new malaria clusters. Descriptive spatial statistics, spatio-temporal analysis, and spatial autocorrelation were performed in order to characterize the geographic features, estimate spatial measures of the data, identify and assess malaria clusters, and ultimately determine whether or not the spatial clustering of malaria incidence across study areas and time periods is due to chance. ArcGIS V10.8 software was used to visualize the results, and SatScan V9.11 was employed for spatiotemporal analysis.

## DESCRIPTIVE SPATIAL STATISTICS

The map showing the incidence rate of malaria in the years 2013 and 2014 is shown in excellent detail in Figure 1. In LGAs including Iwajowa, Kajola, Ibarapa East, Akinyele, Ido, Ibarapa Central, Ibarapa East, Olorunsogo, and Egbeda, fewer instances of malaria were reported in 2014 as compared to 2013. Malaria incidences increased in areas like as Iseyin, Saki East, Atiba, Oyo West, Ogbomoso North, Ogbomoso South, Surulere, Afijio, Ogo Oluwa, Lagelu, and Ona Ara. Furthermore, malaria occurrences in LGAs such Saki West, Irepo, Atisbo, Ibarapa North, and Itesiwaju did not significantly increase or decrease during either time period, indicating little improvement in those areas. Overall, more people have contracted malaria in 2014 than in 2013. This might be a result of factors like population growth, an increase in malaria testing, or subpar disease management.

**Figure 1:**
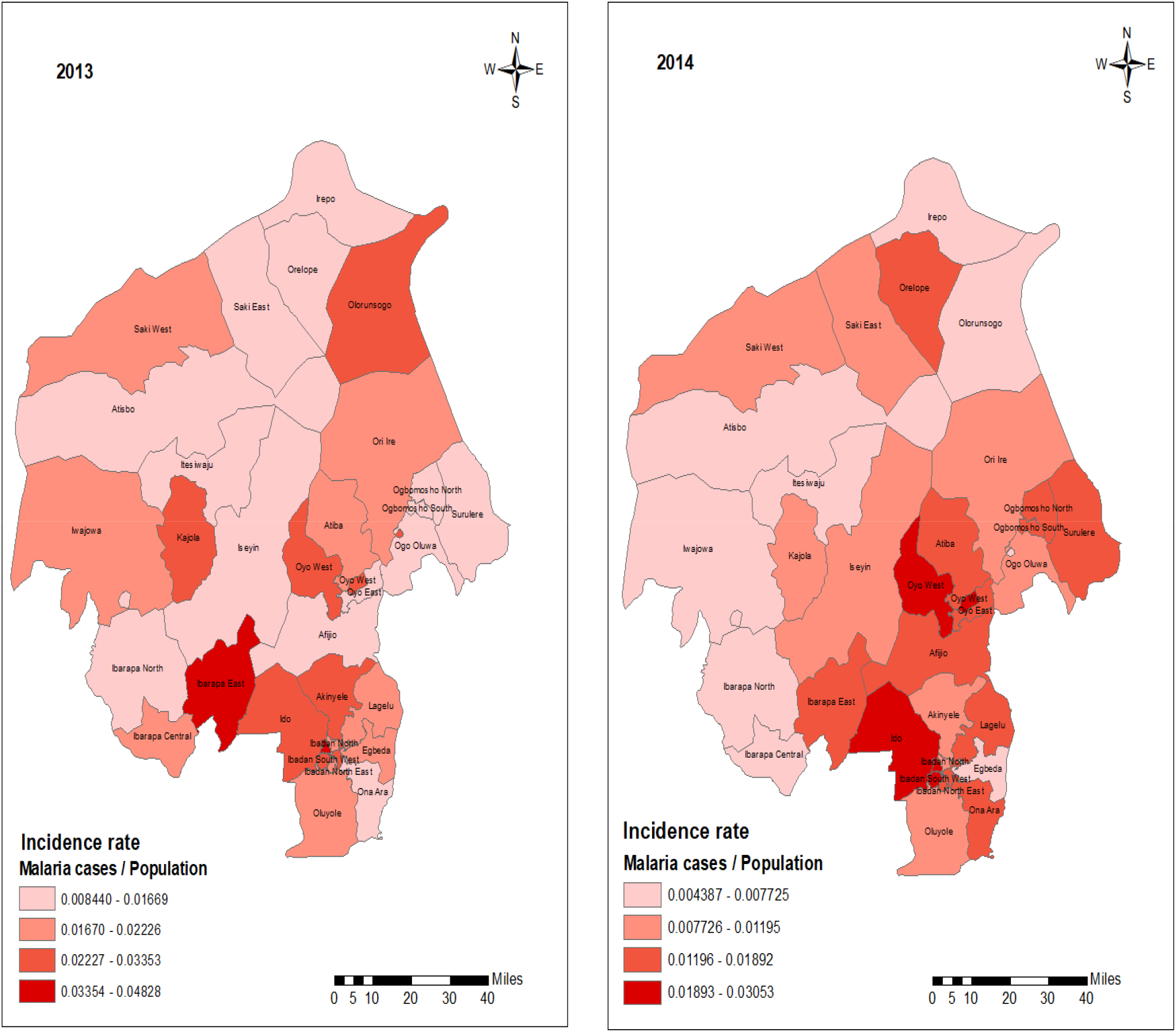
Spatial distribution of malaria incidence by LGAs in Oyo state, 2013-2014.

Figure 2 below clearly depicts the map displaying the incidence rate of malaria in the years 2015 and 2016. Less cases of malaria were reported in 2016 compared to 2015 in LGAs such Irepo, Oluyole, Orelope, Kajola, Itesiwaju, Olorunsogo, Atisbo, Iwajowa, Iseyin, Afijio, Ido, Akinyele, and Lagelu. Atiba and Ori Ire saw an increase in malaria cases, whilst Saki West, Saki East, Ogbomoso North, and Surulere LGAs saw no improvement. Due to the fact that there were fewer malaria cases reported in 2016, there was an improvement from 2015 to 2016.

**Figure 2:**
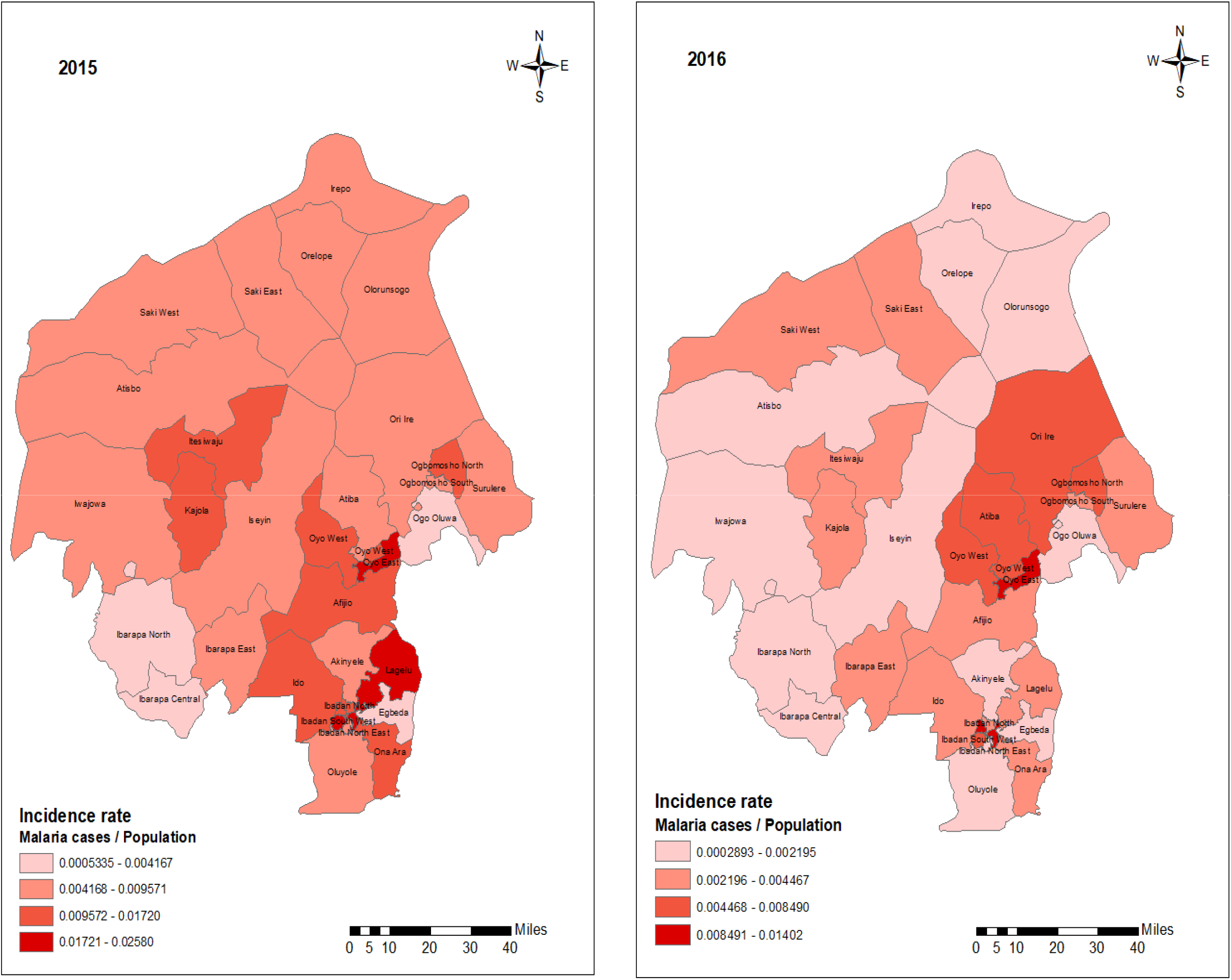
Spatial distribution of malaria incidence by LGAs in Oyo state, 2015-2016.

Figure 3 below makes it evident that from 2013 to 2016, Oyo South reported the largest number of malaria cases, while Oyo North recorded the lowest. Four LGAs (Ibarapa East, Ibadan North, Ibadan North East, and Ibadan South West) out of the nine LGAs in Oyo South contributed more than 50% of the total malaria cases reported in that region from 2013 to 2016.

**Figure 3:**
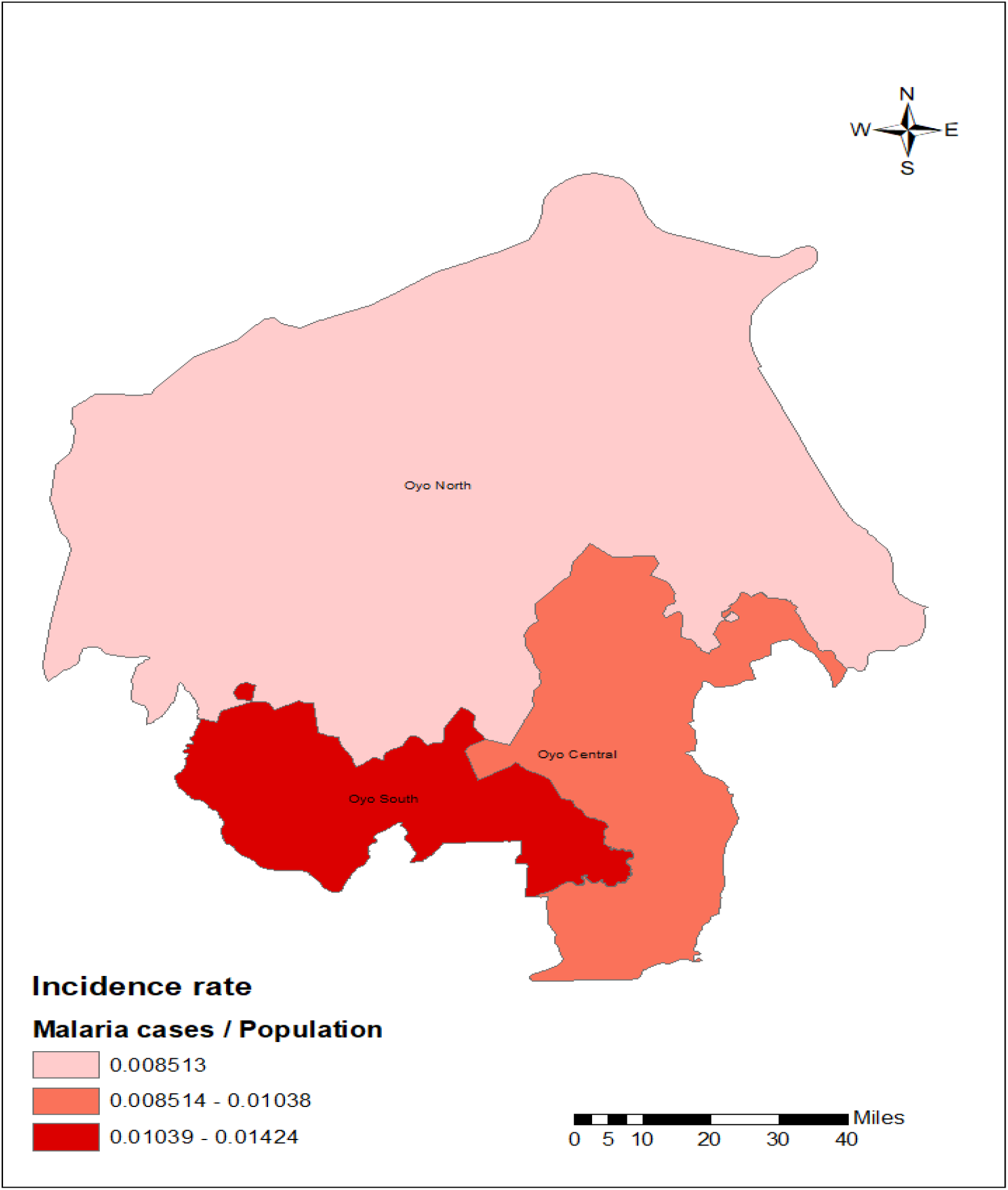
Spatial distribution of malaria incidence by senatorial districts in Oyo state, 2013-2016.

## ESTIMATE MEASURES OF SPATIAL STATISTICS

Table 2 below shows the malaria incidence mean center, Euclidean median, and standard deviation. The mean center and euclidean median are represented by the latitude and longitude 3.79293^0^ and 3.841332^0^ and 7.8999831^0^ and 7.801207^0^, respectively. This implies that the malaria incidence centers in Oyo State are concentrated in the LGAs of Oyo West and Afijio. Additionally, latitude 3.79293^0^ and longitude 7.8999831^0^ are the locations of a standard distance of 0.600391^0^. The standard distance deviation shows that the spread of malaria incidence from Oyo West LGA also covers areas such as Akinyele, Atiba, Afijio, Saki East, Ori Ire, Olorunsogo, Surelere, Atisbo, Iwajowa, Kajola, Iseyin, Itesiwaju, Ogbomoso North, Ogbomoso South, Ibarapa North, Ibarapa Central, Ibarapa East, Ido, Ibadan North, Ibadan North East, Ibadan South East, Ibadan South West, Ibadan North West, Oyo West, Oyo East, Lagelu, Egbeda, Oluyole, Ona Ara and Ogo Oluwa.

**Table 1:** Distribution of the local governments in Oyo state by senatorial districts.

| Oyo state Senatorial Districts | Local Government Areas | Count |
| --- | --- | --- |
| Oyo North | Saki West, Saki East, Orelope, Irepo, Olorunsogo, Ori Ire, Surelere, Atisbo, Iwajowa, Kajola, Iseyin, Itesiwaju, Ogbomoso North, Ogbomoso South. | 14 |
| Oyo Central | Oyo West, Atiba, Oyo East, Afijio, Akinyele, Lagelu, Egbeda, Oluyole, Ona Ara, Ogo Oluwa | 10 |
| Oyo South | Ibarapa North, Ibarapa Central, Ibarapa East, Ido, Ibadan North, Ibadan North East, Ibadan South East, Ibadan South West, Ibadan North West. | 9 |

**Table 2:** Spatial measures.

| **Spatial Measures** | **Statistic** | **Latitude** | **Longitude** | **Location** |
| --- | --- | --- | --- | --- |
| Mean Center | 22367.727 | $3.79293^0$ | $7.8999831^0$ | Oyo west |
| Euclidean Median | 18961 | $3.841332^0$ | $7.801207^0$ | Afijio |
| Standard Distance | $0.600391^0$ | $3.79293^0$ | $7.8999831^0$ | Thirty LGAs |

**Source:** Computed by the author

**Table 2:**
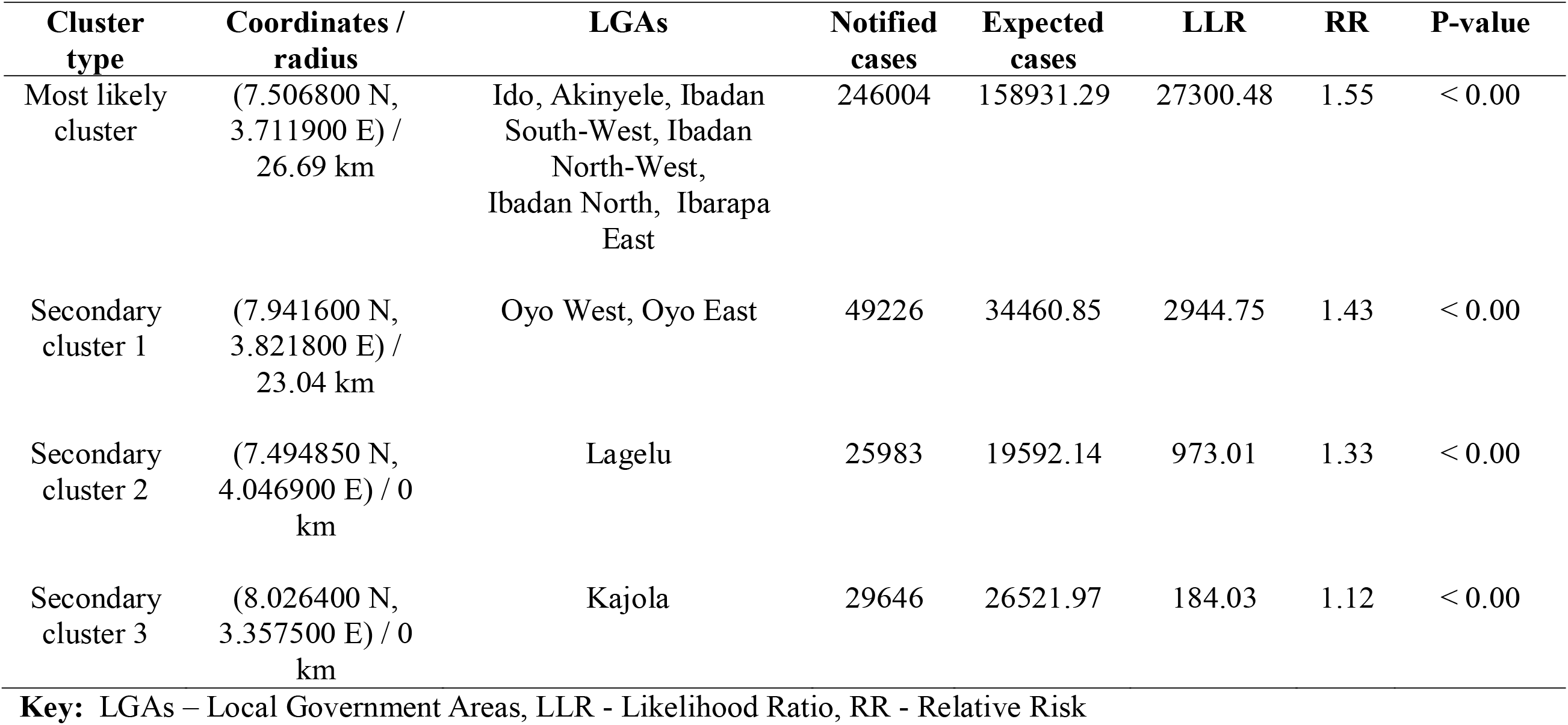
Spatial analysis results of malaria incidence in Oyo state.

| Cluster type | Coordinates / radius | LGAs | Notified cases | Expected cases | LLR | RR | P-value |
| --- | --- | --- | --- | --- | --- | --- | --- |
| Most likely cluster | (7.506800 N, 3.711900 E) / 26.69 km | Ido, Akinyele, Ibadan South-West, Ibadan North-West, Ibadan North, Ibarapa East | 246004 | 158931.29 | 27300.48 | 1.55 | < 0.00 |
| Secondary cluster 1 | (7.941600 N, 3.821800 E) / 23.04 km | Oyo West, Oyo East | 49226 | 34460.85 | 2944.75 | 1.43 | < 0.00 |
| Secondary cluster 2 | (7.494850 N, 4.046900 E) / 0 km | Lagelu | 25983 | 19592.14 | 973.01 | 1.33 | < 0.00 |
| Secondary cluster 3 | (8.026400 N, 3.357500 E) / 0 km | Kajola | 29646 | 26521.97 | 184.03 | 1.12 | < 0.00 |

**Key:** LGAs – Local Government Areas, LLR - Likelihood Ratio, RR - Relative Risk

## SPATIAL ANALYSIS OF MALARIA INCIDENCE

***H***_**0 :**_ Malaria cluster is not statistically significant over space

***H***_**1 :**_Malaria cluster is statistically significant over space

Table 2 below makes it evident that four significant malaria clusters were identified at the 5% level. Ido, Akinyele, Ibadan South West, Ibadan North –West, Ibadan North, and Ibarapa East LGAs have the most probable cluster (RR = 1.55). Oyo West and Oyo East LGA are home to the first secondary cluster (RR = 1.43). Lagelu LGA is home to the second secondary cluster (RR = 1.33), and Kajola is home to the third secondary cluster (RR = 1.12). As a result, the null hypothesis is rejected at the 5% significance level, and it is determined that there is enough data to draw the conclusion that malaria clusters are significant everywhere.

## SPATIO-TEMPORAL ANALYSIS OF MALARIA INCIDENCE

***H***_**0 :**_ Malaria cluster is not statistically significant over space and time

***H***_**1 :**_Malaria cluster is statistically significant over space and time

Table 3 below makes it evident that three substantial malaria clusters were identified at the 5% level. Between January 2013 and December 2014, the Ido, Akinyele, Ibadan South West, Ibadan North –West, Ibadan North, and Ibarapa East LGAs had the highest probability of clustering (RR = 2.27). The Oyo south region is mostly home to the most likely cluster. The first secondary cluster was discovered between January 2013 and December 2014 at Atisbo, Saki West, Saki East, kajola, Itesiwaju, and Iwajowa (RR = 1.75). The second secondary cluster was discovered between January 2013 and December 2014 at Ogo Oluwa, Ogbomoso South, Ogbomoso North, Atiba, Oyo East, Oyo West, Surulere, Ori Iri, Afijio, and Lagelu LGA (RR = 1.48). Hence, the null hypothesis is rejected at 5% level of significance and it is concluded there is sufficient evidence that malaria cluster are significant across the geographical locations and time periods.

**Table 3:** Spatio-temporal results of malaria incidence in Oyo state, 2013–2016.

| Cluster type | Coordinate s / radius | Cluster time | LGAs | Notified cases | Expected cases | LLR | RR | P-value |
| --- | --- | --- | --- | --- | --- | --- | --- | --- |
| Most likely cluster | (7.506800 N, 3.711900 E) / 26.69 km | 2013/1/1 - 2014/12/31 | Ido, Akinyele, Ibadan South-West, Ibadan North-West, Ibadan North, Ibarapa East | 142397 | 62686.82 | 42022.06 | 2.27 | < 0.00 |
| Secondary cluster 1 | (8.406210 N, 3.254728 E) / 48.92 km | 2013/1/1 - 2013/12/31 | Atisbo, Saki West, Saki East, Kajola, Itesiwaju, Iwajowa | 38133 | 21850.69 | 5138.43 | 1.75 | < 0.00 |
| Secondary cluster 2 | (8.406210 N, 3.254728 E) / 48.92 km | 2013/1/1 - 2014/12/31 | Ogo Oluwa, Ogbomosho South, Oyo East, Atiba, Ogbomosho North, Surulere, Oyo West, Ori Ire, Afijio, Lagelu | 105439 | 71120.02 | 8097.50 | 1.48 | < 0.00 |

**Key:** LGAs – Local Government Areas, LLR - Likelihood Ratio, RR - Relative Risk

## SPATIAL AUTOCORRELATION (ANSELIN LOCAL MORAN I)

***H***_**0 :**_Spatial pattern of Malaria incidence is random across the study area

***H***_**1 :**_ Spatial pattern of Malaria incidence is not random across the study area

Table 4 below shows that at the 5% level, Akinyele, Egbeda, Ibadan North, Ibadan North East, Ibadan North West, Ibadan South East, Ibadan South West, and Oluyole have a considerable High High concentration of malaria incidence. The low to high clustering of malaria incidence in these areas is indicated by the Low High concentration of malaria incidence observed in LGAs like Lagelu and Ido. Another sign of high to low malaria clustering in this area is the high-low concentration in Atiba. There is a strong likelihood that LGAs with high-high and low-high concentrations of malaria incidence will impact their neighbors with low or no clustering. Incidence of malaria is seen in very low concentrations in Ogbomoso North, Ogo Oluwa, and Surulere. Since P values < 0.05 level of significance, the null hypothesis of randomness is therefore rejected and it is concluded that spatial pattern is clustered, which further depicts strong spatial dependence across study area in year 2013. The geographic visual representation of the local Moran I is displayed in figure 6.

**Table 4:** Local Moran I dependence of malaria incidence among Local governments areas (2013)

| **LGAs** | **LMI Index** | **LMI Z score** | **LMI P value** | **Concentration** |
| --- | --- | --- | --- | --- |
| Akinyele | 0.000401 | 3.21 | 0.05 | HH |
| Atiba | -0.000015 | - 1.69 | 0.036 | HL |
| Egbeda | 0.00045 | 3.71 | 0.002 | HH |
| Ibadan North | 0.003638 | 4.16 | 0.002 | HH |
| Ibadan North East | 0.001904 | 4.46 | 0.002 | HH |
| Ibadan North West | 0.003044 | 4.54 | 0.002 | HH |
| Ibadan South East | 0.001544 | 3.85 | 0.002 | HH |
| Ibadan South West | 0.002376 | 3.69 | 0.002 | HH |
| Ido | -0.000012 | - 4.42 | 0.002 | LH |
| Lagelu | -0.000111 | -3.62 | 0.002 | LH |
| Ogbomoso North | 0.000092 | 1.87 | 0.016 | LL |
| Ogo Oluwa | 0.000191 | 1.68 | 0.026 | LL |
| Oluyole | 0.000163 | 3.68 | 0.002 | HH |
| Ona Ara | -0.000013 | -3.96 | 0.002 | LH |
| Surulere | 0.000082 | 1.60 | 0.038 | LL |

**Key:** HH – High High, HL – High Low, LL – Low Low, LH – Low High

**Figure 4:**
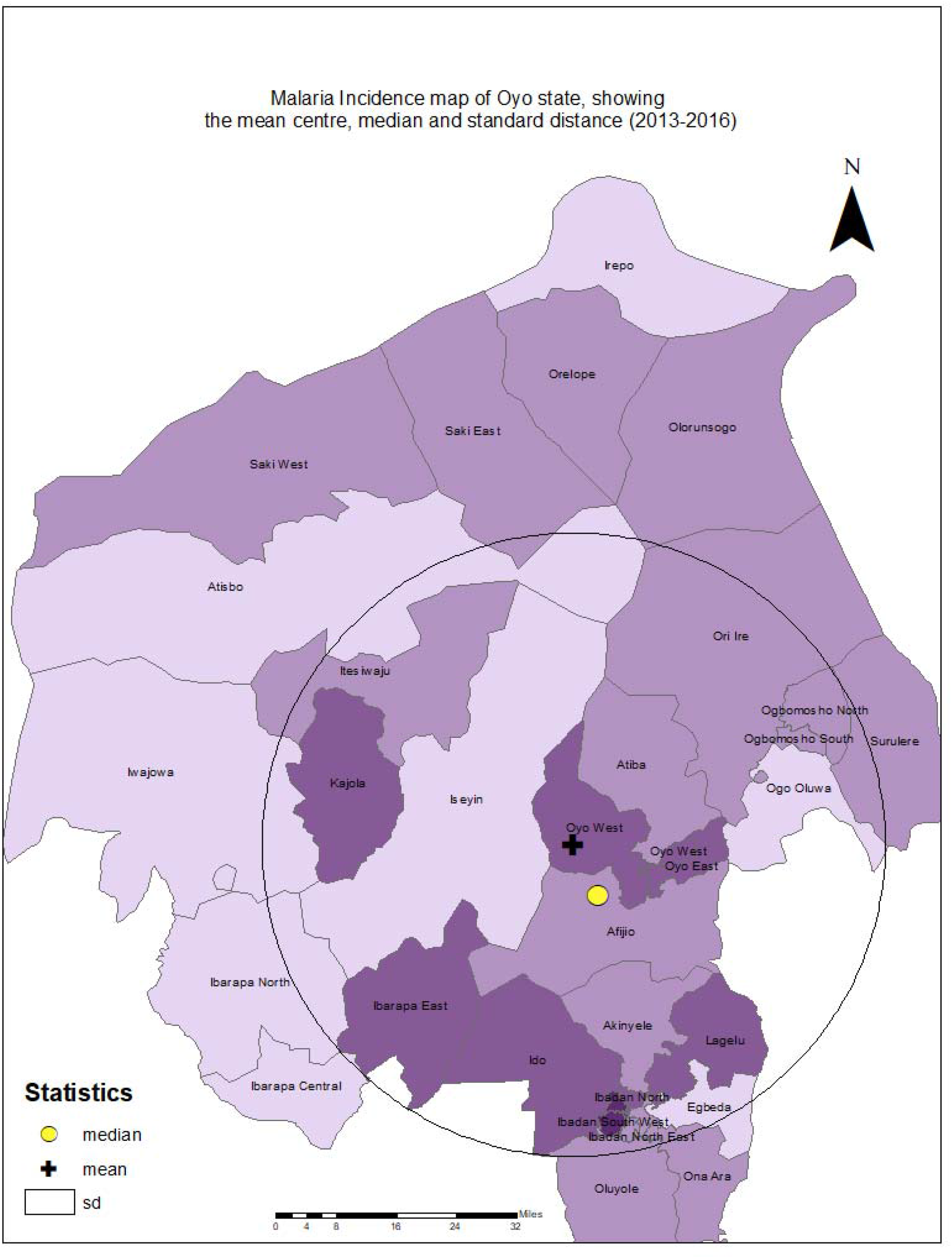
Map showing the Mean center, Euclidean median and Standard distance of malaria incidence in Oyo state, 2013 – 2016.

**Figure 5:**
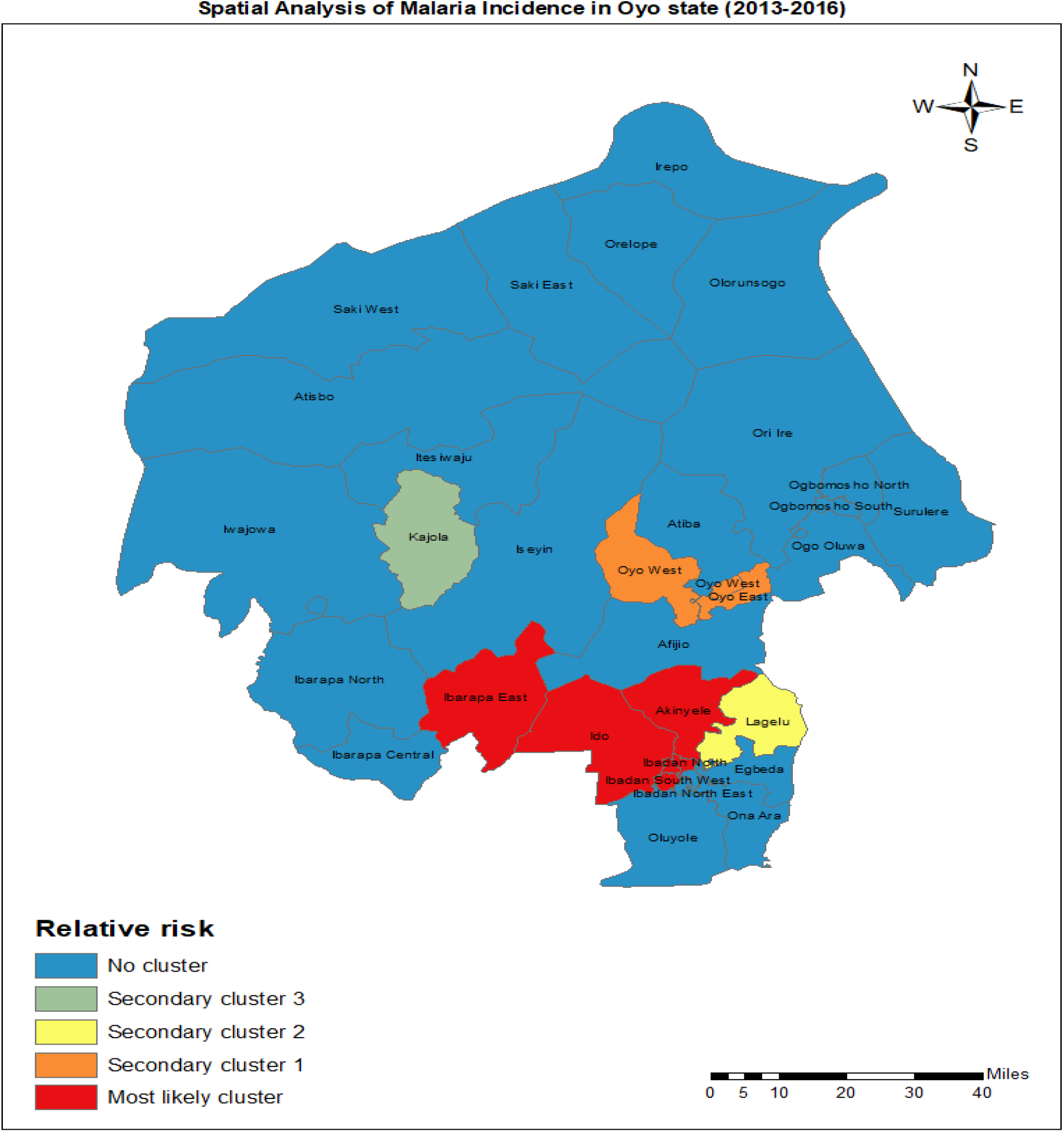
Spatial Analysis of Malaria incidence in Oyo state, 2013-2016.

**Figure 6:**
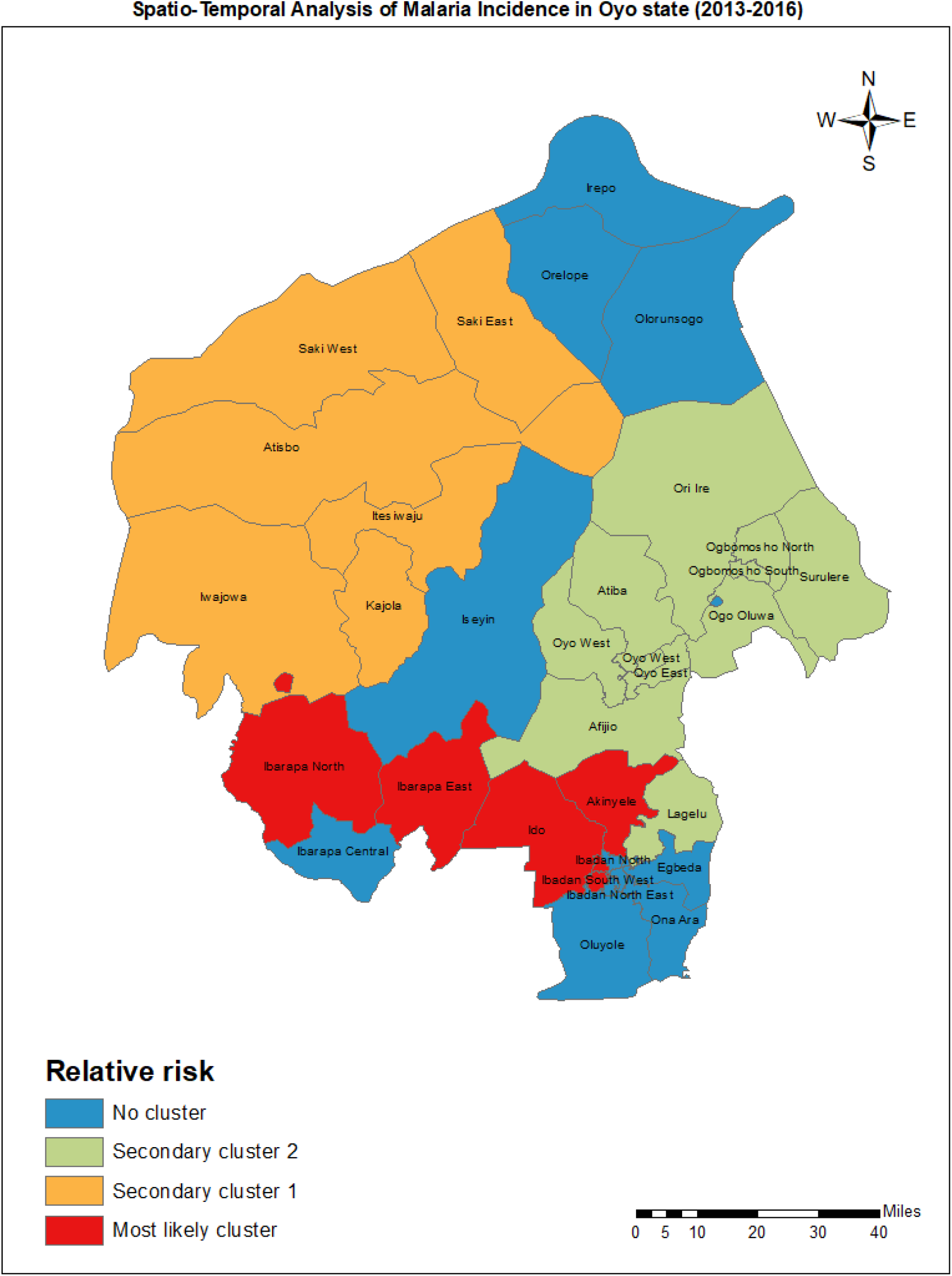
Spatio-Temporal Analysis of Malaria incidence in Oyo state, 2013-2016.

**Figure 6:**
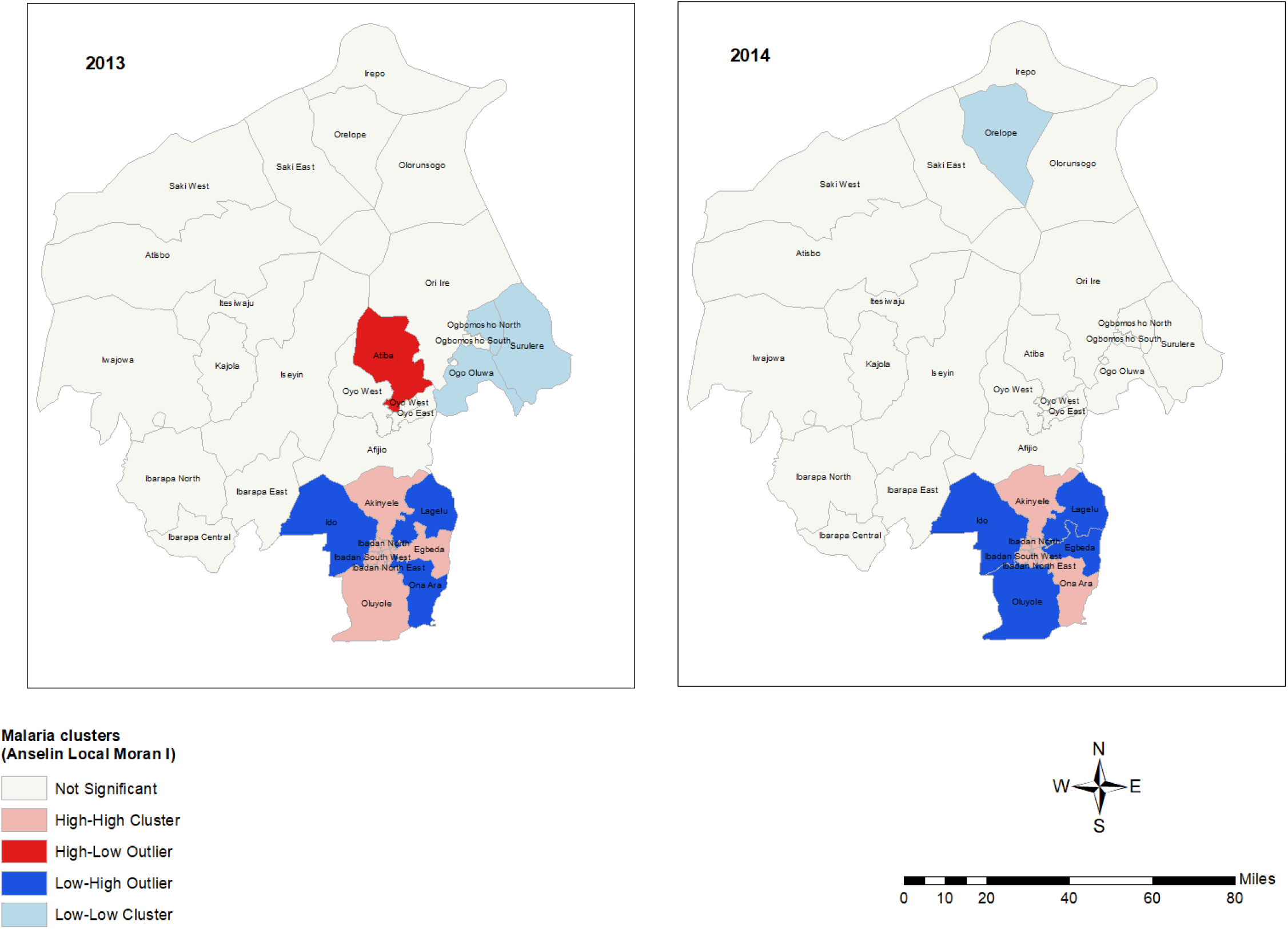
Spatial Autocorrelation of Malaria Incidence in Oyo state, 2013-2014 (Anselin Local Moran I)

Table 5 below shows that Akinyele, Ibadan North, Ibadan North East, Ibadan North West, Ibadan South East, Ibadan South West, and Ona Ara have significant High High concentrations of malaria incidence at the 5% threshold. Low to high clustering of malaria incidence in these locations is indicated by the Low High concentration of malaria incidence observed in LGAs such Lagelu, Ido, Oluyole, and Egbeda. There is a strong likelihood that LGAs with high-high and low-high concentrations of malaria incidence will impact their neighbors with low-low or no clustering. Surulere exhibits a very low concentration of malaria incidence. Since P values < 0.05 level of significance, the null hypothesis of randomness is therefore rejected and it is concluded that spatial pattern is clustered, which further depicts strong spatial dependence across study area in year 2014. The geographic visual representation of the local Moran I is displayed in figure 6.

**Table 5:** Local Moran I dependence of malaria incidence among Local governments areas (2014)

| **LGAs** | **LMI Index** | **LMI Z score** | **LMI P value** | **Concentration** |
| --- | --- | --- | --- | --- |
| Akinyele | 0.000008 | 3.21 | 0.002 | HH |
| Egbeda | -0.000342 | -3.68 | 0.004 | LH |
| Ibadan North | 0.001208 | 3.83 | 0.002 | HH |
| Ibadan North East | 0.001916 | 3.76 | 0.002 | HH |
| Ibadan North West | 0.002213 | 4.21 | 0.002 | HH |
| Ibadan South East | 0.002508 | 4.48 | 0.002 | HH |
| Ibadan South West | 0.003782 | 4.06 | 0.04 | HH |
| Ido | -0.00001 | - 3.41 | 0.004 | LH |
| Lagelu | -0.000016 | -2.83 | 0.008 | LH |
| Oluyole | -0.000021 | -4.42 | 0.002 | LH |
| Ona Ara | 0.0006 | 3.16 | 0.006 | HH |
| Surulere | 0.000044 | 1.68 | 0.016 | LL |

**Key:** HH – High High, HL – High Low, LL – Low Low, LH – Low High

Table 6 below shows that at the 5% level, Ibadan North, Ibadan North East, Ibadan North West, Ibadan South West, Lagelu, and Ona Ara have a statistically significant High High concentration of malaria incidence. In LGAs like Akinyele, Egbeda, Ibadan South East, Ido, and Oluyole, there is a low to high concentration of malaria incidence, which suggests that malaria prevalence is clustered in these regions. There is a strong likelihood that LGAs with high-high and low-high concentrations of malaria incidence will impact their neighbors with low-low or no clustering. The locations of Ibarapa Central, Ibarapa East, and Ibarapa North exhibit a very low concentration of malaria prevalence. Since P values < 0.05 level of significance, the null hypothesis of randomness is therefore rejected and it is concluded that spatial pattern is clustered, which further depicts strong spatial dependence across study area in year 2015. The geographic visual representation of the local Moran I is displayed in figure 7.

**Table 6:** Local Moran I dependence of malaria incidence among Local governments areas (2015)

| **LGAs** | **LMI Index** | **LMI Z score** | **LMI P value** | **Concentration** |
| --- | --- | --- | --- | --- |
| Akinyele | -0.000113 | -3.22 | 0.002 | LH |
| Egbeda | -0.000428 | -3.80 | 0.002 | LH |
| Ibadan North | 0.0001344 | 3.19 | 0.01 | HH |
| Ibadan North East | 0.002356 | 2.71 | 0.018 | HH |
| Ibadan North West | 0.000723 | 3.86 | 0.002 | HH |
| Ibadan South East | -0.000462 | -4.79 | 0.002 | LH |
| Ibadan South West | 0.002204 | 2.28 | 0.03 | HH |
| Ibarapa Central | 0.000058 | 1.15 | 0.028 | LL |
| Ibarapa East | 0.000042 | 1.32 | 0.038 | LL |
| Ibarapa North | 0.000052 | 1.26 | 0.012 | LL |
| Ido | -0.000082 | -3.09 | 0.006 | LH |
| Lagelu | 0.000284 | 2.58 | 0.006 | HH |
| Ogbomoso North | -0.000065 | -1.18 | 0.04 | HL |
| Oluyole | -0.000152 | -3.43 | 0.002 | LH |
| Ona Ara | 0.000345 | 2.72 | 0.006 | HH |

**Key:** HH – High High, HL – High Low, LL – Low Low, LH – Low High

**Figure 7:**
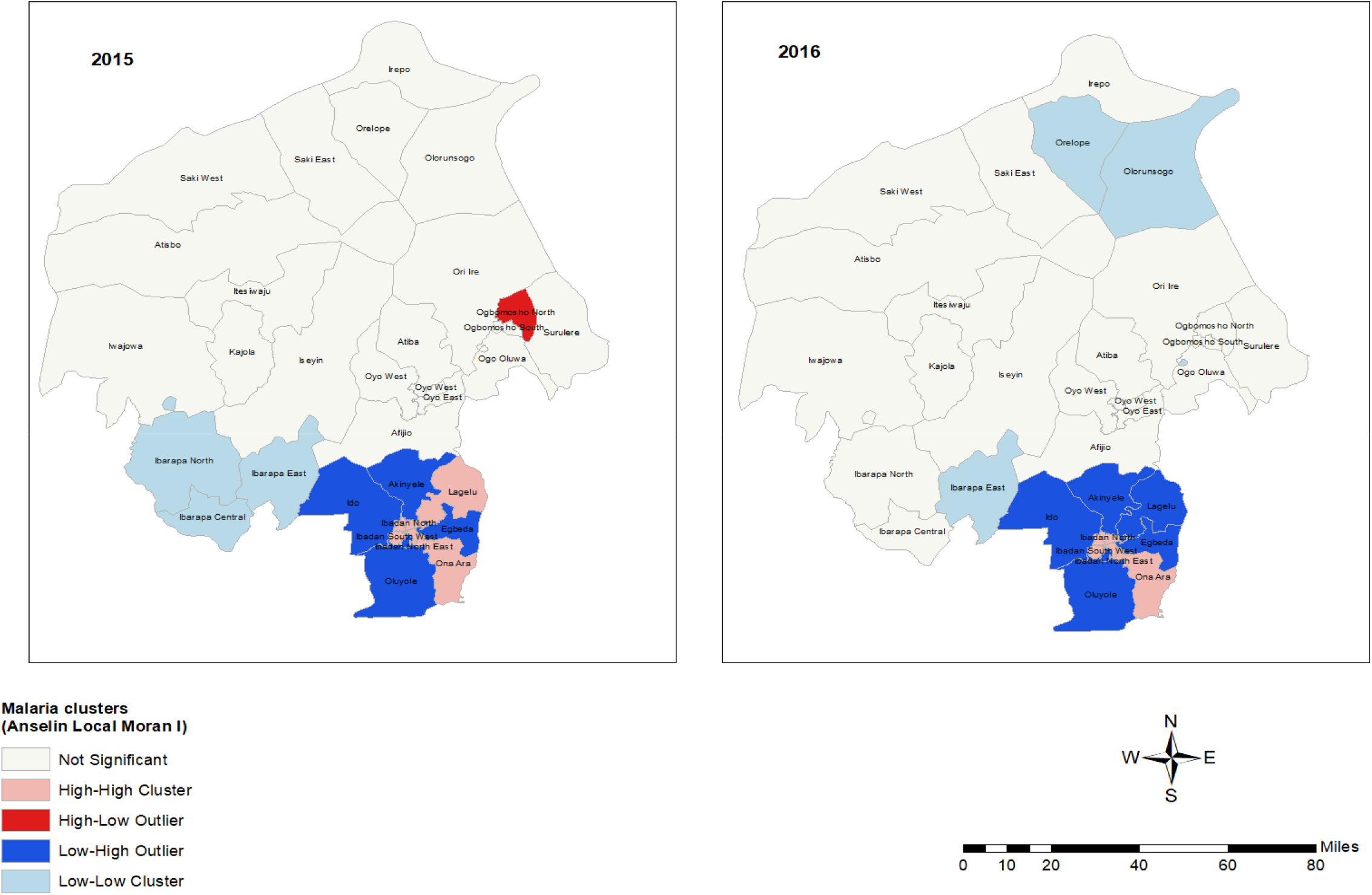
Spatial Autocorrelation of Malaria Incidence in Oyo state, 2015 - 2016 (Anselin Local Moran I)

Table 7 below shows that at the 5% level, Ibadan North, Ibadan North East, Ibadan North West, Ibadan South West, and Ona Ara have a statistically significant High High concentration of malaria incidence. The low to high clustering of malaria incidence in these locations is indicated by the Low High concentration of malaria incidence observed in LGAs such Akinyele, Egbeda, Ibadan South East, Ido, Lagelu, and Oluyole. There is a strong likelihood that LGAs with high-high and low-high concentrations of malaria incidence will impact their neighbors with low-low or no clustering. Ibarapa East, Irepo, Olorunsogo, and Surulere exhibit a very low concentration of malaria incidence. Since P values < 0.05 level of significance, the null hypothesis of randomness is therefore rejected and it is concluded that spatial pattern is clustered, which further depicts strong spatial dependence across study area in year 2016. The geographic visual representation of the local Moran I is displayed in figure 7.

**Table 7:** Local Moran I dependence of malaria incidence among Local governments areas (2016)

| **LGAs** | **LMI Index** | **LMI Z score** | **LMI P value** | **Concentration** |
| --- | --- | --- | --- | --- |
| Akinyele | -0.000141 | -2.84 | 0.004 | LH |
| Egbeda | -0.000227 | -2.80 | 0.006 | LH |
| Ibadan North | 0.001266 | 3.91 | 0.004 | HH |
| Ibadan North East | 0.00229 | 2.50 | 0.026 | HH |
| Ibadan North West | 0.001618 | 3.13 | 0.006 | HH |
| Ibadan South East | -0.000266 | -4.75 | 0.002 | LH |
| Ibadan South West | 0.001971 | 2.52 | 0.028 | HH |
| Ibarapa East | 0.000021 | 1.38 | 0.02 | LL |
| Ido | -0.000173 | -2.50 | 0.016 | LH |
| Irepo | 0.000042 | 1.13 | 0.046 | LL |
| Lagelu | -0.000039 | -2.40 | 0.01 | LH |
| Olorunsogo | 0.000035 | 1.26 | 0.032 | LL |
| Oluyole | -0.000276 | -3.23 | 0.002 | LH |
| Ona Ara | 0.000033 | 2.54 | 0.02 | HH |
| Surulere | 0.000053 | 1.32 | 0.048 | LL |

**Key:** HH – High High, HL – High Low, LL – Low Low, LH – Low High

## SPATIAL AUTOCORRELATION (GLOBAL MORAN I)

Table 8 below displays the 2013 Moran I index calculation result, which is 0.479. The Z score is 4.95 standard deviations, and the p value is 0.001. According to this result, there is a less than 5% chance that the clustered pattern is not a coincidental coincidence. For 2014, the calculated value of the Moran I index is 0.371. The Z score is 4.02 standard deviations, and the p value is 0.002. According to this result, there is a less than 5% chance that the clustered pattern is not a coincidental coincidence. The outcome of the 2015 Moran I index computation was 0.088. The Z score is 1.23 standard deviations, and the p value is 0.114. This result implies that the likelihood that the clustered pattern is a random chance is more than 5%. The outcome of the 2016 Moran I index computation was 0.129. The Z score is 1.71 standard deviations, and the p value is 0.061. This conclusion implies that the likelihood that the clustered pattern is the product of chance is larger than 5%. From 2013 to 2016, the computed value of the Moran I index is 0.273. The Z score is 11.67 standard deviations, and the p value is 0.001. According to this result, there is a less than 5% chance that the clustered pattern is not a coincidental coincidence.

**Table 8:** Moran I Global Spatial Autocorrelation results.

| **Time period** | **Moran's I** | **Z score** | **P-value** |
| --- | --- | --- | --- |
| 2013 | 0.479 | 4.9542 | 0.001 |
| 2014 | 0.371 | 4.0236 | 0.002 |
| 2015 | 0.088 | 1.2332 | 0.114 |
| 2016 | 0.129 | 1.7124 | 0.061 |
| 2013 – 2016 | 0.273 | 11.6733 | 0.001 |

**Source:** Computed by author

## CONCLUSION

The results of the study corroborate the hypothesis that spatial analyses identified four major malaria clusters. The most likely cluster is concentrated in the Oyo South region. Spatial-temporal studies also revealed three main malaria clusters. The most likely cluster, which is thought to have formed between January 2013 and December 2014, is located in the Oyo South and Oyo Central region. Six local government units make up the first secondary cluster in Oyo North. In all the geographical and spatio-temporal studies, the same LGAs—Ido, Akinyele, Ibadan South West, Ibadan North -West, Ibadan North, and Ibarapa East—are located in the most likely cluster. The spatial dependence of malaria incidence distribution is illustrated by the findings of the local moran I spatial autocorrelation, which showed a considerable clustering of malaria cases throughout the research area.

## Data Availability

All data produced in the present study are available upon reasonable request to the authors

## APPENDIX

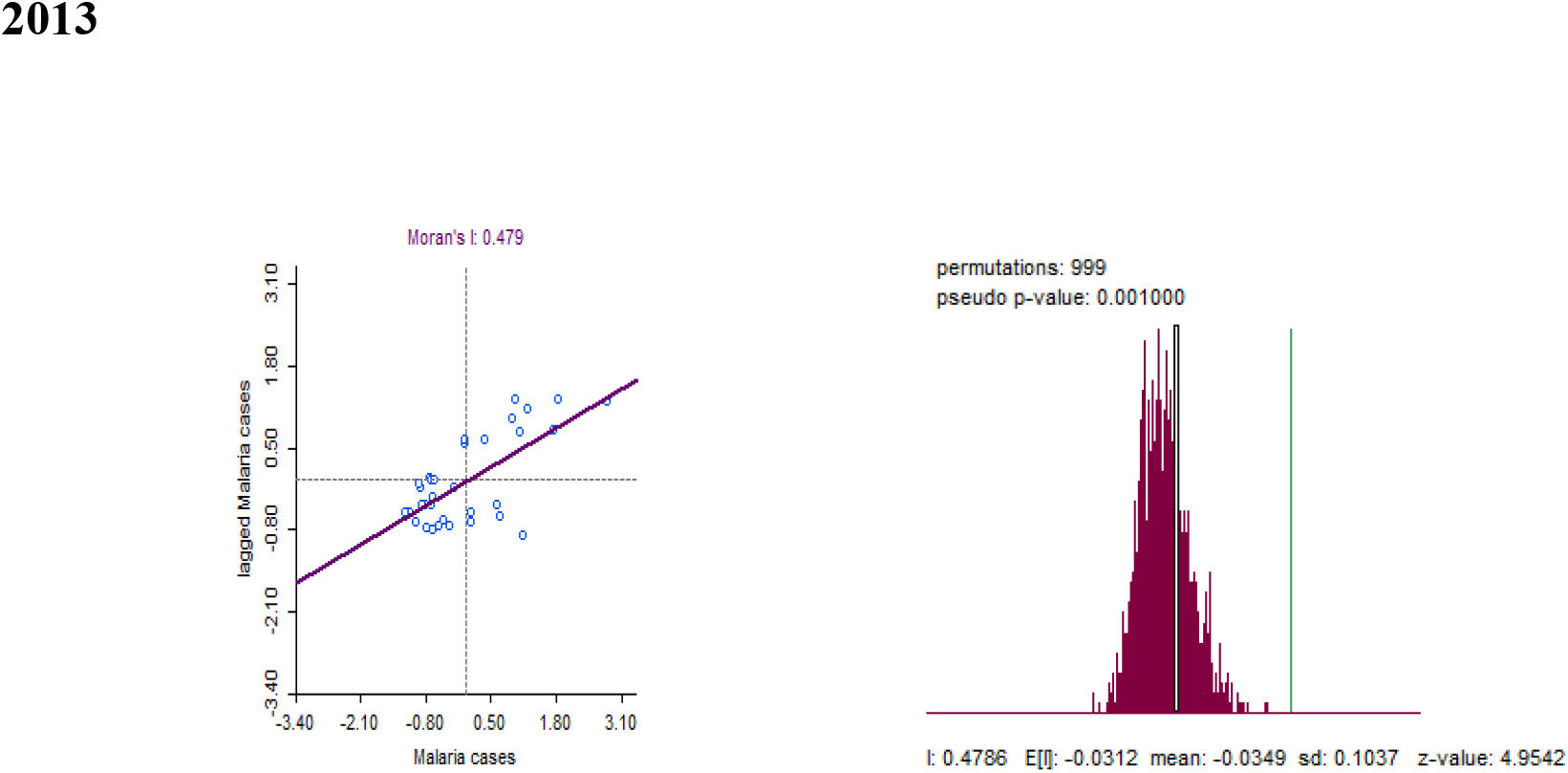

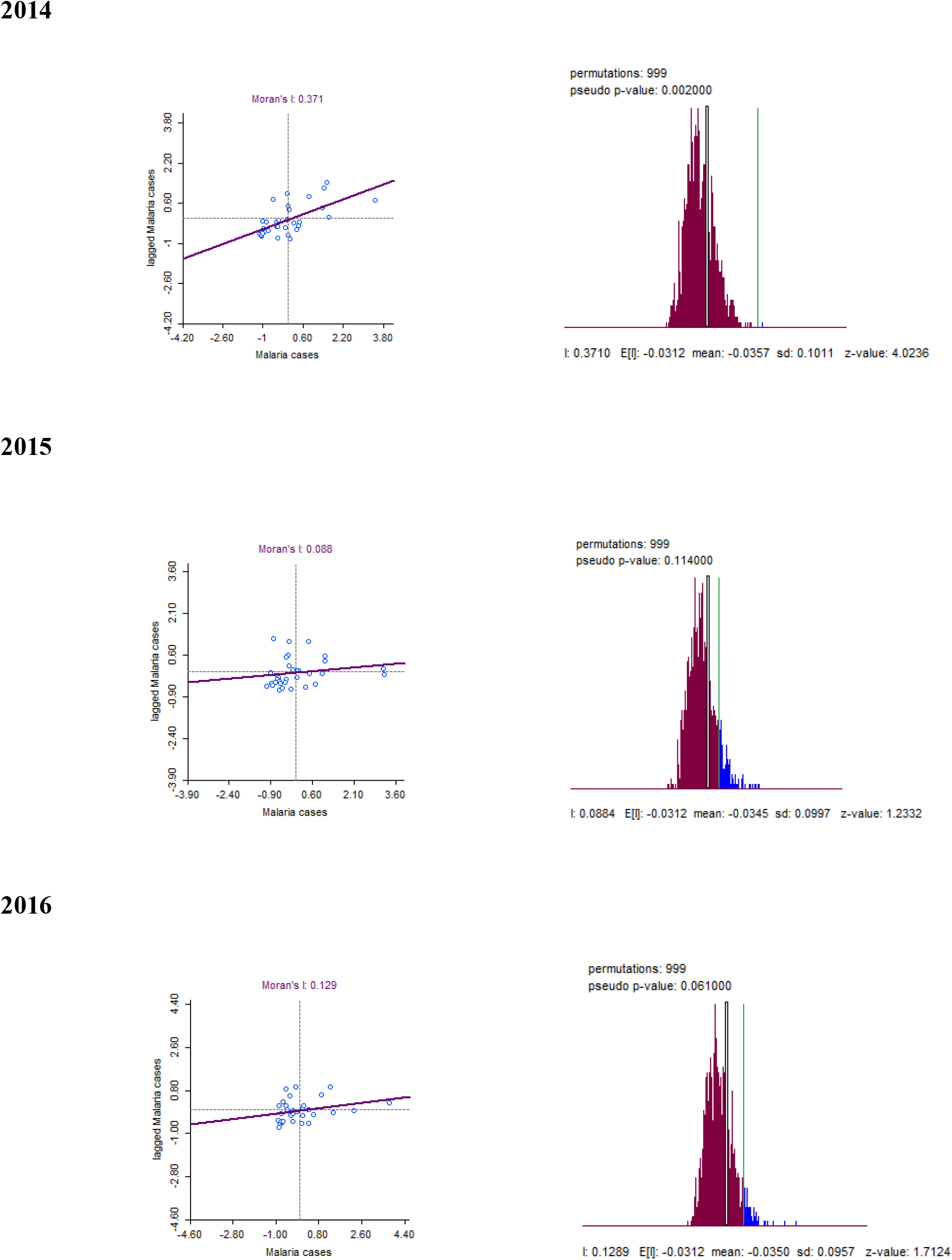

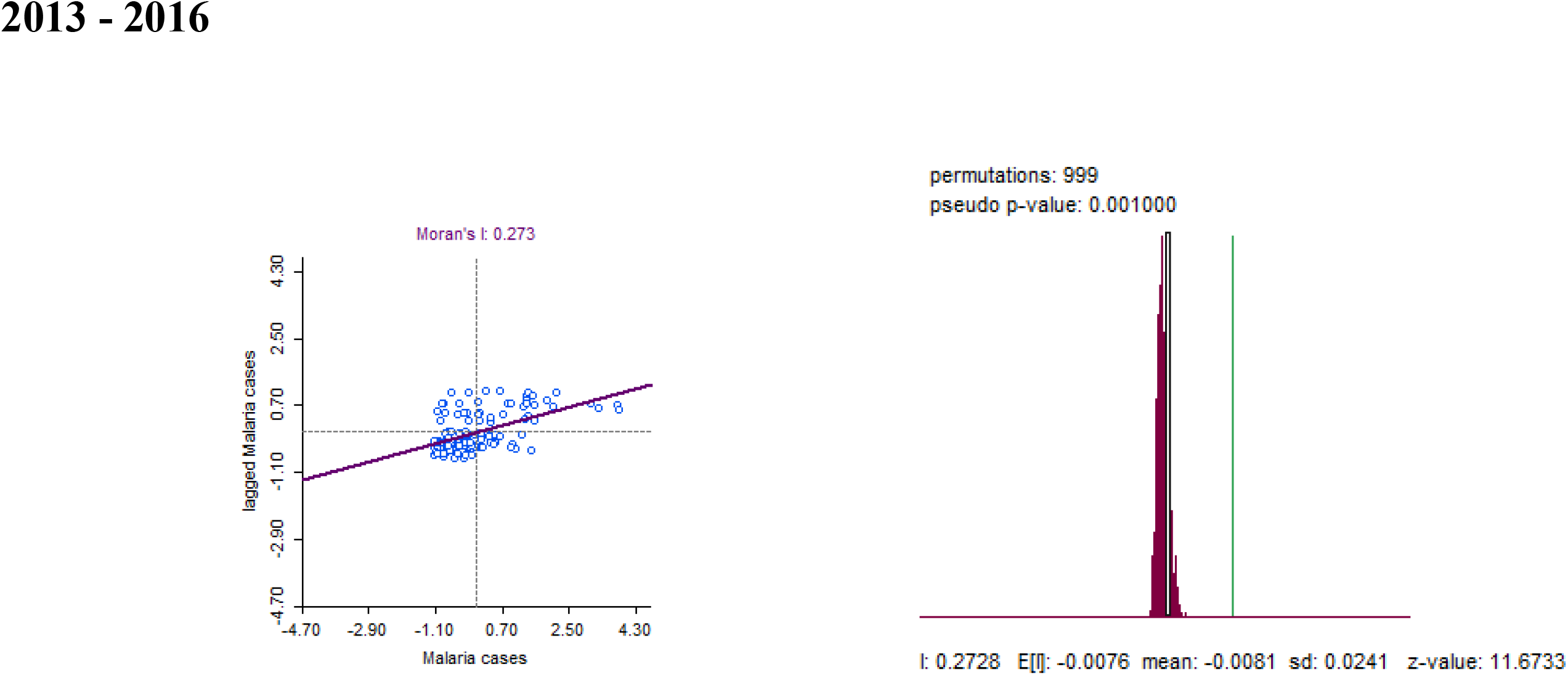

**GLOBAL MORAN I**

Note. The image in figure 1 was sourced from Tulane University website. https://www2.tulane.edu/~wiser/protozoology/notes/images/mal_lc.gif. The image in figure 2 was sourced from Krekora et al. (2017); the course of pregnancy and delivery in a patient with malaria. Ginekologia Polska. 88. 574-575. 10.5603/GP.a2017.0103.

